# Erythrovirus B19 (B19V) in patients with acute febrile illness suspected of arboviruses in Mato Grosso do Sul, Brazil

**DOI:** 10.1101/2023.11.29.23299091

**Authors:** Gislene Garcia C. Lichs, Zoraida del Carmen Fernandez Grillo, Valdinete Alves do Nascimento, Daniel Maximo Corrêa Alcantara, Everton Ferreira Lemos, Cristiano M. Espínola Carvalho, Luiz Henrique Ferraz Demarchi, Crhistinne Carvalho Maymone Gonçalves, Felipe Gomes Naveca, Alexsandra Rodrigues de Mendonça Favacho

## Abstract

Human Erythrovirus (parvovirus) B19 infection can produce symptoms similar to those produced by Dengue, Chikungunya, and Zika viruses, making clinical diagnosis difficult. The importance of erythrovirus B19 in human pathology has been increased and reported in numerous studies published globally. The B19V infection was investigated by real-time PCR in samples from patients with signs and symptoms related to classic arboviral symptoms. This study was conducted to provide information on the genetic diversity of Human Erythrovirus B19 (B19V) circulating in the state of Mato Grosso do Sul, Midwest region of Brazil, from 2017 to 2022. A total of 773 sera samples of patients with negative diagnostic results for Dengue, Chikungunya, and Zika, during the study period were analyzed. Erythrovirus DNA was found in 10.6% (82/773) of patients, among them 10 were pregnant women. Four samples were completely sequenced, and the other five partially, to genotype by phylogenetic reconstruction. All samples belong to worldwide dispersed genotype 1, subgenotype 1a. These results demonstrate the importance of including B19V in differential laboratory diagnosis for epidemiological purposes and appropriate patient management. The diagnosis for B19V should be performed, particularly among pregnant women, immunocompromised patients, and individuals with hemolytic diseases, as the infection is more severe in these cases.

## Introduction

Human Erythrovirus B19 (parvovirus B19), of the *Parvoviridae* family, a member of the *Erythroparvovirus* genus, was discovered in England by Yvonne Cossart et al in 1975 [1] in serum samples from blood donors who performed serological testing for hepatitis B virus. This sample was coded as “number 19 in panel B”, and later the virus was called B19 [1].

After its discovery, the B19V infection was associated with an asymptomatic or benign acute pediatric infection known as erythema infectiosum and other clinical manifestations such as transient aplastic crisis, and arthropathies [2].

The transmission of B19V is through the respiratory droplets. However, the virus can also be transmitted parenterally, especially by haemoderivative applications [3]. The Erythrovirus B19 has a tropism for human erythroid progenitor cells and it is involved in suppressing blood cell formation during infection [4]. In 2002, the virus was classified into three genotypes, defined as genotype 1 B19 classic (represented by the prototype strain Au), genotype 2 (prototype K71 and strain A6-similar), and genotype 3 (prototype V9) [5].

The infection by Human Erythrovirus B19 during pregnancy has been widely studied, and it is known to cause a range of complications, including spontaneous abortion [2]. The virus acts on the inhibition of red blood cell formation, generating cytotoxic effects that lead to variable clinical conditions, such as intrauterine growth retardation, myocarditis, and pericardial effusions, which will depend on the patient’s hematological and immunological status. The age group most affected is children/adolescents under 14 years old [6]. However, many infections are asymptomatic and are not correctly diagnosed.

In Brazil, the B19V and some arboviruses (Dengue, Chikungunya, and Zika) co-circulate in several regions of the country, making the differential diagnosis difficult, mainly because these viruses can cause similar symptoms associated with exanthema and acute febrile diseases [7].

The disease caused by B19V was not classified as national or state compulsory notification, in Brazil. The infection was detected in surveillance of exanthematous diseases, as a differential diagnosis in suspected cases of measles or rubella, according to the characteristics of symptoms [8]. In 2020, in a meeting between the State Secretary of Health of Mato Grosso do Sul (SES) and the Central Laboratory of Public Health (LACEN) it was agreed that suspected cases of Erythema Infectiosum (B19V) should be notified, but it was only from 2021 that some cases were included in the Notification Diseases Information System (SINAN). The detection of IgM antibodies against parvovirus B19 is carried out at the LACEN following national guidelines and protocols provided by the Ministry of Health.

Diagnosis for B19V, using real-time PCR, should be performed particularly among pregnant women, immunocompromised patients, and individuals with hemolytic diseases since the infection is more serious in these cases. The implementation in the laboratory routine can clarify whether erythrovirus B19 may be one of the etiological agents of exanthematous diseases, as well as determine the frequency of their infection in patients.

The current study aims to evaluate the presence of Human Erythrovirus B19 in samples from patients with negative diagnostic results for Dengue, Chikungunya, and Zika collected in several municipalities in the state of Mato Grosso do Sul, Brazil, from 2017 to 2022, and provide information of molecular characterization and genetic variability of circulating strains of Human Erythrovirus B19 in the state.

## Materials and Methods

### Study design and sampling

Biological samples were selected from the Biobank of the Central Laboratory of Public Health of Mato Grosso do Sul state (LACEN/MS), based on the following criteria: (i) serum, urine, and cerebrospinal fluid samples collected between January 2017 and October 2022; (ii) with clinical suspicion of chikungunya, dengue or zika disease but with negative test results (“not detectable” by the RT-qPCR and “non-reactive” when using the NS1 and enzyme- linked immunosorbent assay (ELISA) for these arboviruses; (iii) reported onset of symptoms within 5 days for the NS1 test; and (iv) availability at the time of sample selection, since a substantial number of samples were discarded during the laboratory routine due to a lack of storage capacity, a condition exacerbated by the SARS-CoV-2 pandemic; v) samples with a volume equal or more than 200 microliters.

Socio-demographic and clinical data of each sample were obtained from the medical request forms to assess associations with Erythrovirus B19 infection, including, gender, age, city of residence, reported symptoms, pregnancy, and confirmation of death. In addition, the records of positive cases of CHIKV, DENV, and ZIKV during the routine diagnosis of LACEN/MS were compared with the qPCR results obtained for B19V in the present study.

### Ethical Approval

This study was conducted with the authorization of the Human Research Ethical Committee of the Dom Bosco Catholic University of Mato Grosso do Sul, Brazil (CAAE:54019821.2.0000.5162).

### Virus detection

The nucleic acids of each sample were extracted using the QIAsymphony DSPVirus/Pathogen Mini Kit (QIAGEN), according to the manufacturer’s protocol. The extracted DNA was tested for B19V using the protocol developed by Naveca et al. (data not yet published) from the Leônidas and Maria Deane Institute (ILMD), Fiocruz Amazônia.

Primers and probes are presented in Table S1. Real-time PCR (qPCR) was performed using the KAPA PROBE FAST qPCR Master Mix (2X) (Roche), following the calculations shown in Table S2. In all the reactions, the amplification of the human internal control gene (RNase P) was used to rule out false negatives, thereby confirming the accuracy of the results. A no- template control and positive control for B19V were used to discard possible contamination and validate the reaction. The qPCR was performed using the ABI 7500 and QuantStudio™ 5 (Applied Biosystems). Following the manufacturer’s protocol, it applied an initial denaturation at 95°C for 5’, with 40 cycles of denaturation at 95°C for 3” and combined annealing/extension/data acquisition at 60°C for 30”. The threshold for the quantification cycle (Cq) was calculated automatically with default settings using equipment software. The results were considered “positive” for B19V when the Cq value was ≤ 37.

### Sequencing

Samples with positive results for B19V were amplified by conventional PCR using three pairs of primers (Table S3), covering almost the entire viral genome. Nine samples were separated for sequencing (Table 2). They were indicative of the first three years of the study and also had a high viral load. PCR reactions were performed using SuperFi II green enzyme (Thermo Fisher Scientific), primers at 0.5μM, and 1μL of viral DNA in 10μL of the final volume. The recommended cycling parameters were as follows, 98°C of initial denaturation for 30”, 35 cycles of 10” denaturation at 98°C, 10” annealing at 55°C, 2’ extension at 72°C, ending with 5’ final extension at 72°C.

At the end of the reactions, each amplicon was subjected to electrophoresis at 80V for 1 h, visualized on a 1% agarose gel stained with GelRed (Biotium), and the GeneRuler 1 kb DNA Ladder (Thermo Fisher Scientific) was used to confirm the expected size of the product.

Subsequently, the PCR products were precipitated using polyethylene glycol 8000 following an adapted protocol. Initially, 20% PEG8000 (Promega) was added at a 1:1 ratio to the tube containing the PCR product, followed by incubation at 37°C for 15 min. After incubation, centrifugation was performed at 13,000 × g for 15 min. The supernatant was then discarded, 125 µL of 80% ethanol was added, followed by a new centrifugation step at 12,000 × g for 2 min, and the supernatant was discarded. The microtube was then placed in a Mivac DNA concentrator (Genevc SP Scientific) for 15 min at 37°C. DNA was then resuspended in nuclease-free water and quantified using a microvolume spectrophotometer (Biodrop Duo Biochrom). All amplicons from the same sample were pooled together and quantified using the Qubit 2.0 and dsDNA HS Assay kit (Thermo Fisher Scientific).

Four samples were suitable for whole-genome sequencing. Library preparation was performed using a Nextera XT DNA Library Prep Kit (Illumina). Enzymatic tagmentation, adapter, and index addition, amplification, normalization, and pool libraries were performed as described in the manufacturer’s manual using 1ng of DNA. All libraries were quantified using a Qubit dsDNA HS Assay Kit. Sequencing was performed on MiSeq equipment (Illumina) using the Reagent v2 500-cycle sequencing kit. The data generated after sequencing were analyzed in Geneious Prime 2022.0.1 software for the assembly of contigs of each sample, using the genome NC_000883 as the reference and the BBMap v 38.84 tool under “default” conditions. The remaining five samples were partially sequenced (ranging from 585 to 1281 base pairs of the VP1/VP2 gene) using standard procedures for capillary sequencing, as recommended for ABI3130 (ThermoFisher). Trace files were edited for quality and primer removal and assembled using the B19V. The nucleotide sequences obtained during this study were deposited in the GenBank database (Table 2).

### Phylogenetic analysis

The consensus B19V sequences obtained in this study and the reference sequences available on GenBank [G1a: M13178 (isolated Au); G1b: DQ357064 (isolated Vn147); G2: AY064475 (isolated A6); G2 AY044266 (isolated LaLi); G3a: AX003421 (isolated V9) and G3b: AY083234 (isolated D91.1)] were aligned using the MAFFT v7.490 tool with automatic algorithm selection. Subsequently, the alignment file was subjected to maximum-likelihood phylogenetic reconstruction with the FastTree 2.1.11 program using the GTR evolutionary model. An approximate likelihood ratio test (aLRT) [9] was used to assess the branch supports. The phylogenetic tree had the root placed at the central point, with increasing node order, and was edited in the FigTree 1.4.4 program. Tip labels were aligned for clarity.

### Statistical analyses

Mixed Generalized Linear Models (GLMMs) with random effects for municipalities and binomial distribution were used to understand how PCR test results could be related to (1) the age and gender of patients, as well as the year of sampling; and (2) the symptoms recorded in patient records. Symptoms were evaluated in a separate model, owing to fewer records.

Multicollinearity was checked by examining the Pearson correlation coefficient (Pearson’s r) between each pair of explanatory variables, using the R package ‘correlation’ [10], and computing the variance inflation factor (VIF), with the R package ‘performance’ [11]. The analyses were performed using the ‘lme4’ package [12] in the R software [13]. Tables and estimation plots were generated using the ‘jtools’ package [14]. The models were checked for normality of residuals, normality of random effects, homogeneity of variance, and residual dispersion using the packages ‘DHARMa’ [15] and ‘performance’.

In years when sampling was conducted for all months, circular statistics were employed to test seasonal patterns in the number of cases in Mato Grosso do Sul [16–18]. Two approaches were used: (1) a hypothesis test and (2) a model-based approach, both implemented in the R package ‘CircMLE’ [19]. In the hypothesis test, the null hypothesis is that the data are uniformly distributed throughout the year, versus some form of concentration, be it unimodal, bimodal, or multimodal [18]. The Hermans-Rasson test was used to verify the uniformity of the distributions, as it has high statistical power for grouped multimodal distributions [17]. In the model-based approach, the package calculates the maximum likelihood of 10 models described by [20] and compares them using model selection criteria. Thus, this approach allows the identification of patterns of occurrence. Briefly, the models fall into three main categories: (i) a uniform model (M1) of random orientation; (ii) unimodal models (M2A, M2B, M2C) with a single preferred direction; and (iii) bimodal models (M3, M4, and M5) with two preferred directions. Bimodal models can also be divided into axial (M3A, M3B, M4A, M4B) and non-axial (M5A, M5B) [18–21]. Herein, the models were compared based on Akaike’s Information Criterion with small-sample correction (AICc). Additionally, the Akaike weight (*w_i_*) was used, which describes the probability that a particular model is the best model (approximate), given the experimental data and the collection of models considered [21–23]. For each year, it was also calculated the circular standard deviation (sd), circular mean (μ), and the length of the mean vector (r) using the R package ‘circular’ [24]. The μ represents the mean date of cases, and r represents how the data is clustered around the mean (0, perfectly uniformly distributed; and 1, perfectly clustered). All statistical analyses were performed using the R software v4.2.1.

## RESULTS

Between January 2017 and October 2022, LACEN/MS performed 26,154 tests for at least one of the three arboviruses. Some of the samples received for diagnosis of dengue were also tested for CHIKV and/or ZIKV, and others were just evaluated for CHIKV and/or ZIKV. Of the total samples analyzed, 7,239 were tested for CHIKV, 14,247 for DENV, and 4,668 for ZIKV. Of those, 7,162 were negative for CHIKV (98.9%), 7,340 for DENV (51.5%), and 4,634 for ZIKV (99.3%) (Figure 1). Despite the high number of tests performed, few samples were available at the time of sample selection. Therefore, only 773 samples from 62 municipalities in the state of Mato Grosso do Sul met the previously defined criteria, had negative results for dengue, zika, and chikungunya, and were chosen for B19V research (Figure 1). Of the total samples analyzed, 82 were positive for B19V (10.6%), from 24 municipalities (Table 1; Figure 2), surpassing the number of cases registered for Chikungunya and Zika during the same period in the state of Mato Grosso do Sul (Figure 3).

**Figure 1.**
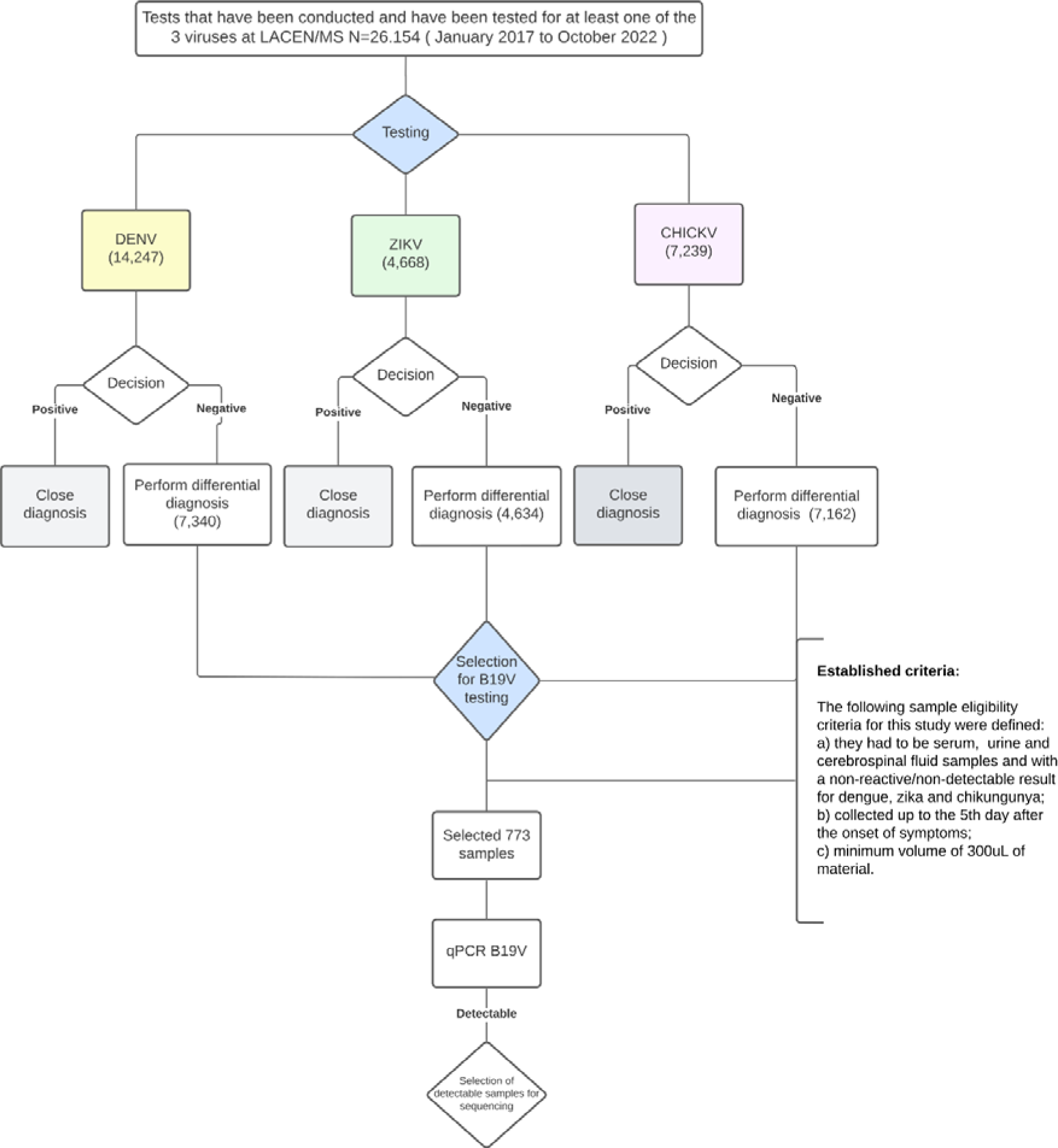
Flowchart of the experimental design of the present study.

**Figure 2.**
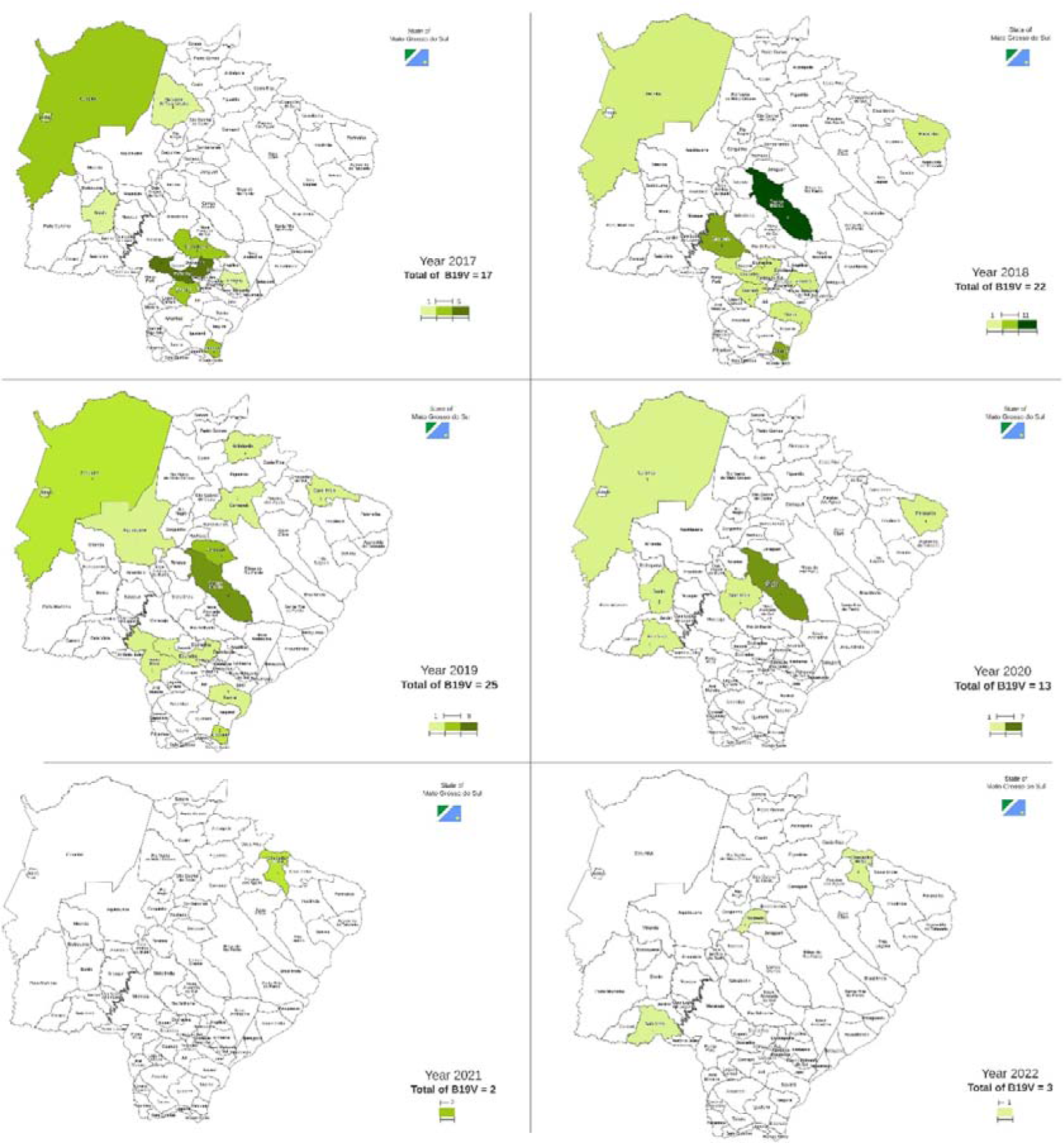
Positive cases of B19V, according to municipalities in the state of Mato Grosso do Sul, during the period from 2017 to 2022.

**Figure 3.**
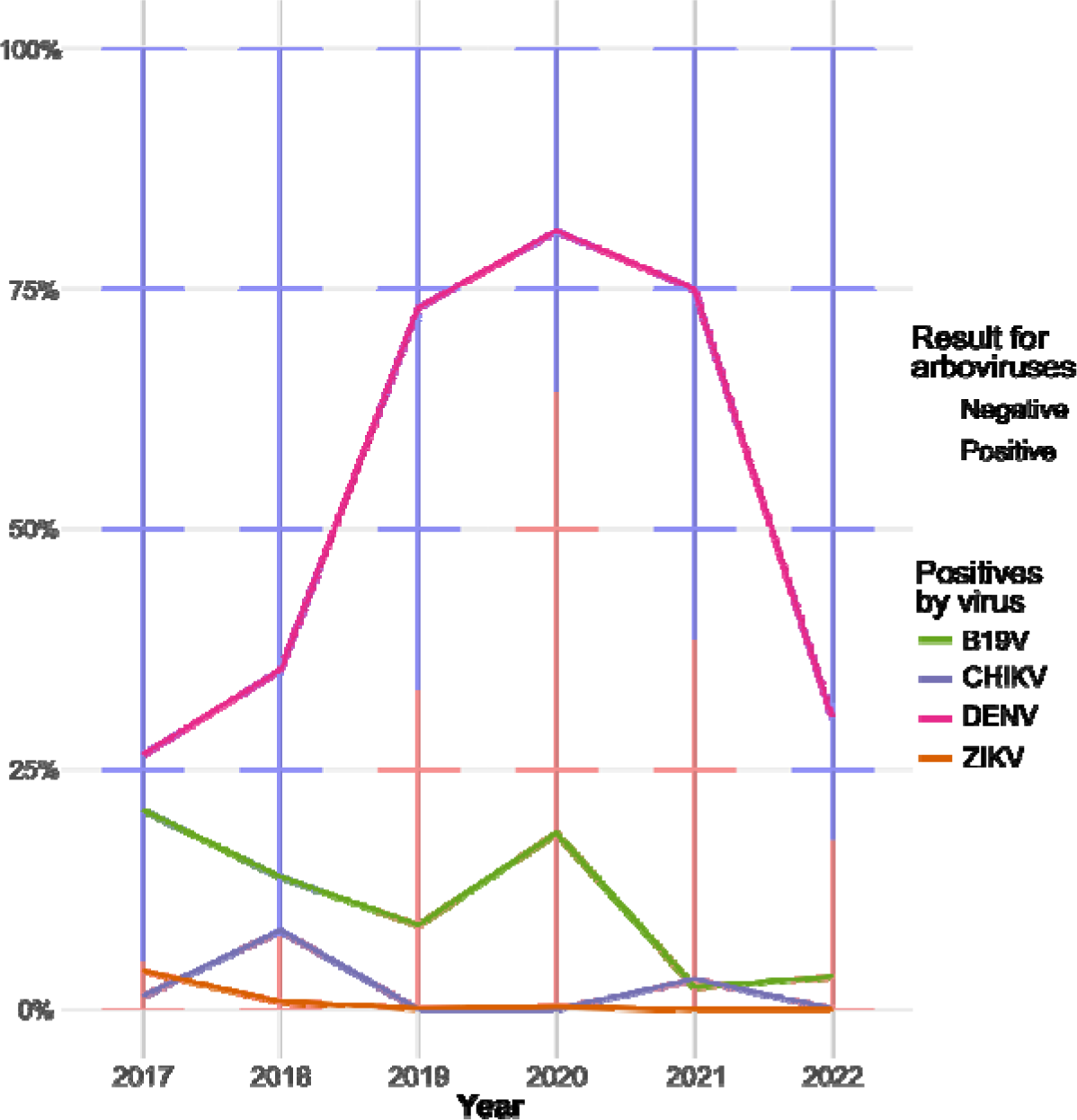
Samples of arboviruses surveyed at LACEN/MS and their results compared to B19V, from 2017 to 2022. The bar graph shows the proportion of molecular test results for all arboviruses examined in LACEN/MS. The lines indicate the percentage of positive results for each virus.

**Table 1:** Population and municipalities studied by year and positive cases for B19V.

| Year | Samples | Positives (%) | Municipalities sampled | Municipalities with positive cases (%) |
| --- | --- | --- | --- | --- |
| 2017 | 82 | 17 (20.7) | 28 | 10 (35.7) |
| 2018 | 160 | 22 (13.8) | 27 | 10 (37.0) |
| 2019 | 285 | 25 (8.8) | 39 | 11 (28.2) |
| 2020 | 71 | 13 (18.3) | 22 | 6 (27.3) |
| 2021 | 86 | 2 (2.3) | 15 | 1 (6.7) |
| 2022 | 89 | 3 (3.4) | 22 | 3 (13.6) |
| Total | 773 | 82 (10.6) | 62 | 24 (38.7) |

Of the total positive samples, 28 were collected in men and 54 in women (Figure S1), with 10 pregnant women. The ages of patients ranged from 1 to 70 years, however, B19V detection was not significantly associated with gender and age group, although it was significantly different between the years of the period evaluated (Figure 4A). The positive results in the years 2019, 2021, and 2022 were significantly lower than in 2017 (Table 1; Figure 4A). The notification forms of 598 patients contained clinical information, identifying a total of 20 symptoms. Among B19V-positive patients, fever was the most frequent symptom, followed by myalgia, headache, arthralgia, retro-orbital pain, nausea, and rash. (Figure 5). Despite this, B19V detection was significantly associated only with retro-orbital pain, leukopenia, petechiae, and malaise (Table 1; Figure 4B). The symptom “abdominal discomfort” was not included in the model because it was correlated with “diarrhea”. The symptoms “gingival bleeding”, “renal failure”, “sore throat” and “thrombocytopenia” were not included because they only had 2 cases in the notification forms, causing problems in the estimates.

**Figure 4.**
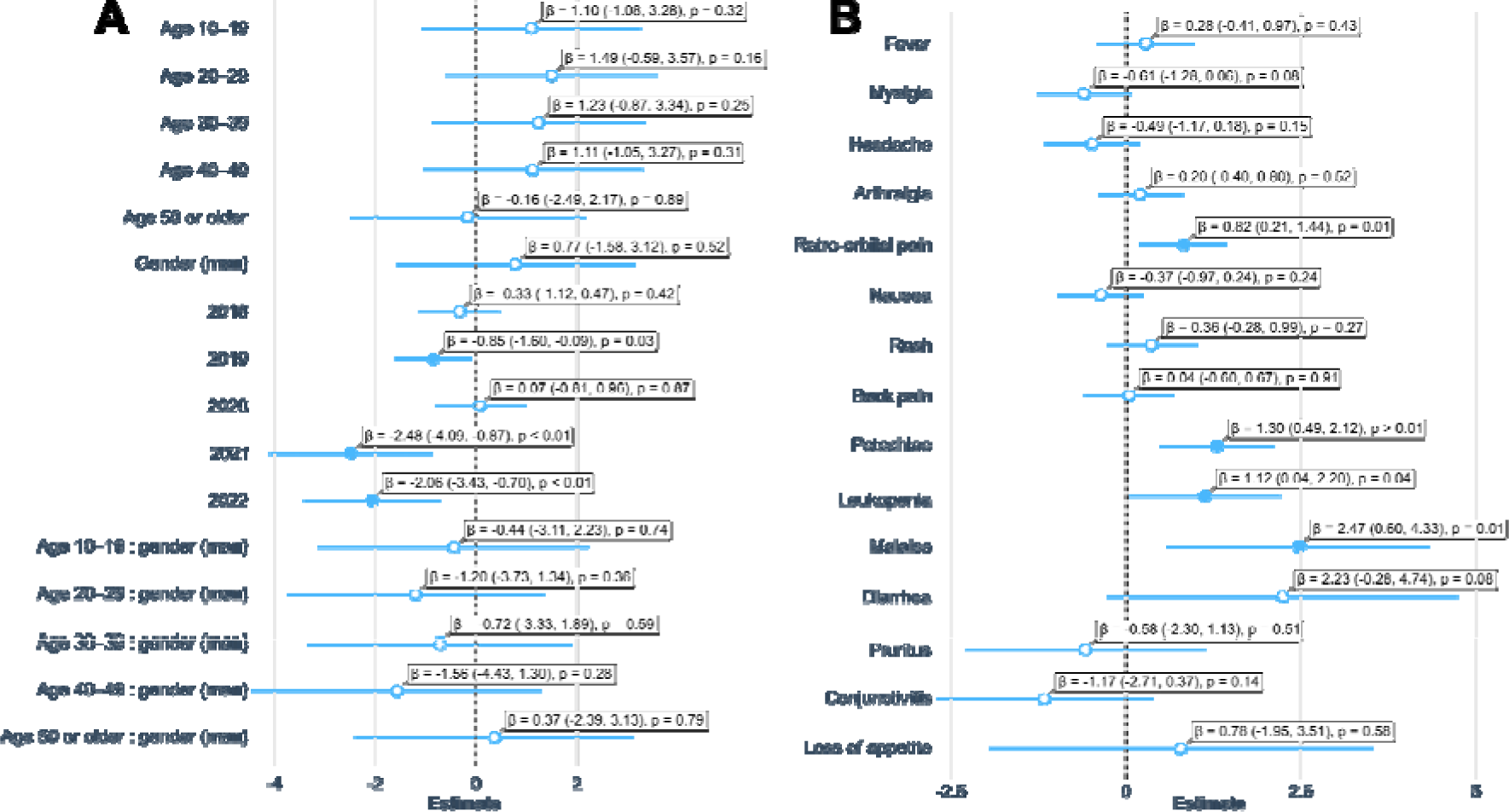
Estimates of mixed generalized linear models. (A) Estimates of patient characteristics and period. (B) Estimates of symptoms. Filled dots indicate significant variables (p < 0.05); bars represent the 95% confidence interval; β, estimate; numbers in parentheses indicate the estimated 95% confidence intervals; p, p-value.

**Figure 5.**
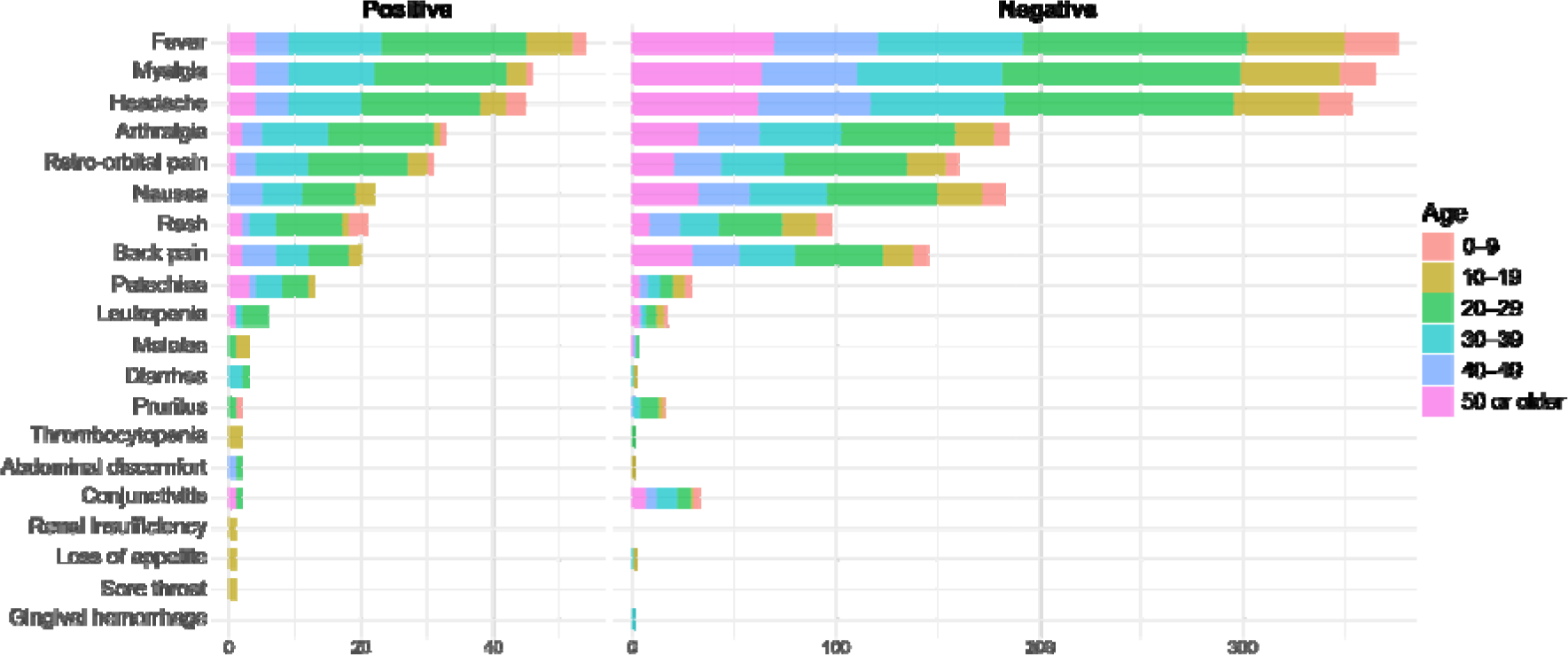
Frequency of symptoms by age group of the studied population.

The phylogenetic reconstruction showed that all samples sequenced in this study were recovered with Genotype 1, subgenotype 1a (RefSeq M13178), with high support (aLRT = 0.94) (Figure 6). The B19V samples that underwent sequencing originate from different municipalities (Corumbá, Campo Grande, Dourados, Deodápolis, Caarapó, Chapadão do Sul, and Alcinópolis) in various geographic areas, demonstrating the wide spread of the virus by the whole state. Identifiers (CodeID) were created for each sample to ensure the confidentiality of the subjects’ identities in the study and to prevent recognition by anyone outside the research group (Table 2; Figure S2).

**Figure 6.**
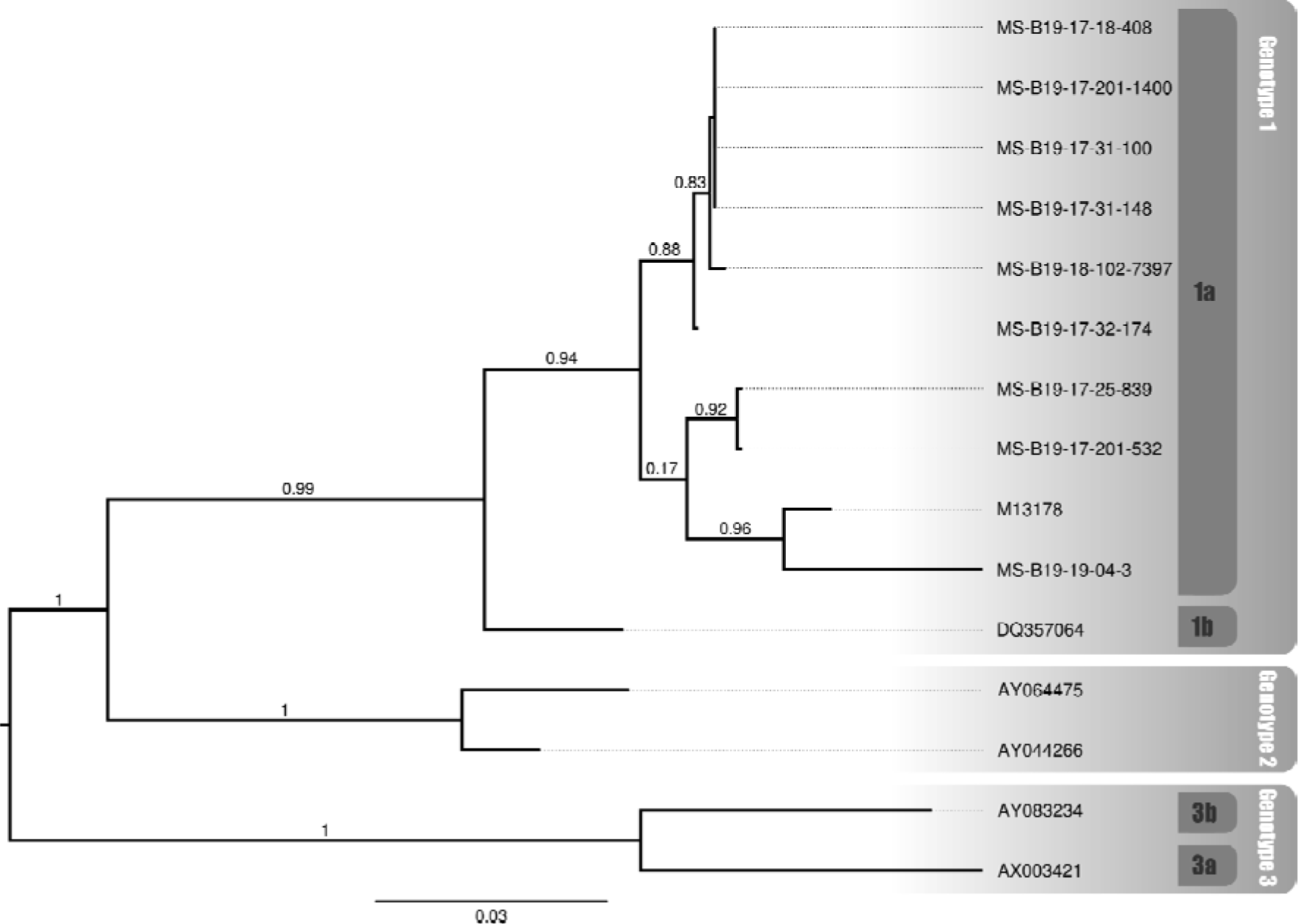
Phylogenetic relationship of sequenced samples and B19V genotypes (1a, 1b, 2, 3a and 3b). The samples sequenced in this study start with “MS-B19”. The reference sequences obtained from GenBank are identified by their accession number. The numbers above branches indicate the approximate likelihood ratio test (aLRT).

**Table 2:** Characteristics of samples sequenced to identify the circulating B19V genotype. All municipalities are in the state of Mato Grosso do Sul, The GenBank access numbers are shown for each sample.

| Code ID | Gender | Municipality | Collection date | Pregnancy | Accession number |
| --- | --- | --- | --- | --- | --- |
| MS-B19-17-201-532 | Woman | Dourados | 03/2017 | Yes | OR542579 |
| MS-B19-17-32-174 | Woman | Chapadão do Sul | 05/2017 | No | OR542583 |
| MS-B19-17-25-839 | Woman | Corumbá | 05/2017 | Yes | OR542582 |
| MS-B19-17-201-1400 | Woman | Dourados | 07/2017 | Yes | OR578525 |
| MS-B19-17-18-408 | Woman | Caarapó | 08/2017 | No | OR542580 |
| MS-B19-17-31-100 | Woman | Eldorado | 10/2017 | No | OR578526 |
| MS-B19-17-31-148 | Man | Eldorado | 11/2017 | - | OR578523 |
| MS-B19-18-102-7397 | Man | Campo Grande | 07/2018 | - | OR578524 |
| MS-B19-19-04-3 | Woman | Alcinópolis | 01/2019 | No | OR542581 |

The results of the Hermans-Rasson test demonstrated that the number of cases was seasonal for all the years evaluated, 2017 (7.29; p = 0.033), 2018 (13.02; p < 0.001), and 2019 (13.90; p < 0.001). For 2017, despite the best model having presented an AICc *w_i_* with more than twice the second, the following models can still be considered plausible choices. When considering the first two models (M3A and M4A, AICc *w_i_* = 0.38 and 0.17; Figure 7; Table S4), they together presented an AICc *w_i_* = 0.55, which means that there is at least a 55% chance that it is the best approximation that describes the data, given the candidate set of orientations considered. Both models are axial bimodal, suggesting a distribution in two equally sized points in 2017, one in each semester, with the mean date of cases in September (mean date = September 01, 2017; sd = 1.67; r = 0.25). In 2018, no model stood out, and the first two were equally supported, with close AICc *w_i_* values, which, together, presented a value of AICc *w_i_* = 0.71 (M5A and M3A, AICc *w_i_* = 0.38 and 0.33; Figure 7; Table S5).

**Figure 7.**
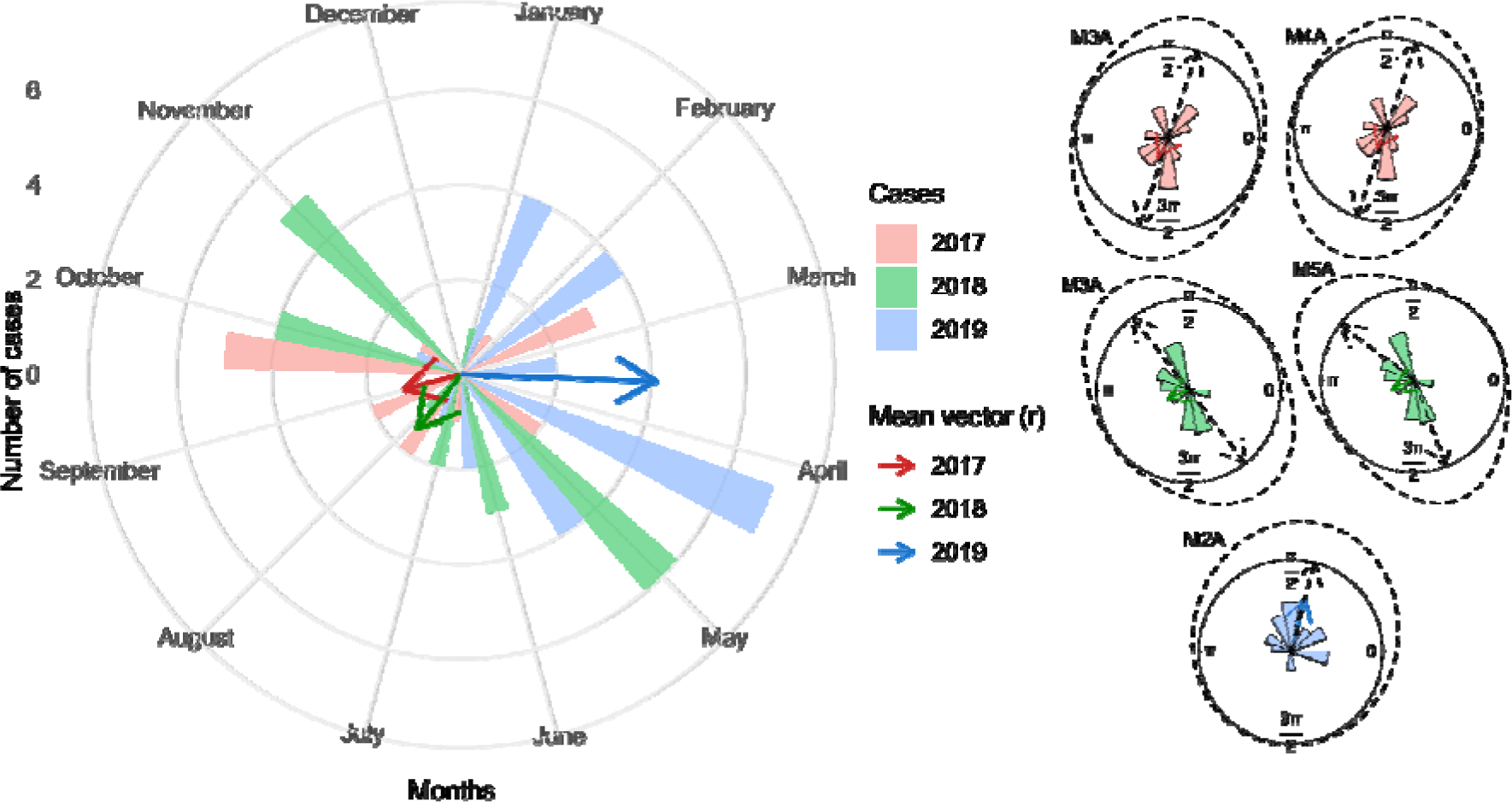
Circular histograms of several B19 cases per month for the years 2017, 2018, and 2019. In the larger histogram, the numbers indicate the number of cases in the respective month, and the mean vector (arrow) is the length and direction of the mean date of cases. The smaller histograms represent the best models found, with the average vector (arrow), the density (dashed line), and the average direction (dashed arrows).

Although both have a bimodal distribution, one is non-axial (M5A), while the other is (M3A), making it more complicated to determine a pattern for the year 2018. Even so, both models showed a division of distribution between the two semesters, with a greater presence of cases closer to the end of each semester, with the mean date of cases in July (mean date = July 23, 2018; sd = 1.68; r = 0.25). In contrast, the pattern shown in 2019 was unequivocal, with an unimodal distribution (M2A, AICc *w_i_* = 0.72; Figure 7; Table S6) concentrated in the first half, with the average date in March (mean date = March 17, 2019; sd = 1.03; r = 0.59).

## DISCUSSION

A wide variety of viruses can cause rashes and joint pain in children, adolescents, and adults, leading to etiological identification exclusively through clinical examination, which makes it a challenging task. The most common agents of exanthematous disease (ED) include Measles, Rubella, Dengue, Chickenpox, Cytomegalovirus, Epstein Barr, Human HerpesVirus 6, Enterovirus, Erythrovirus (Human Parvovirus B19), Chikungunya and Zika [25]. Although these injuries are common in Brazil, there are several difficulties in determining an accurate etiological diagnosis, especially in those regions where there is co-circulation of exanthematous diseases. Therefore, the differential diagnosis of these diseases is extremely important, not only for epidemiological purposes but also for the control and treatment of infections. Even if most of them have a benign course, for certain age groups, pregnant women, and immunocompromised people, some infections represent an important risk, as they can evolve into serious cases, which may require emergency hospitalization, burdening the public health service [6].

Molecular diagnosis of B19V infection is not commonly used as a differential diagnosis, and there are no studies dealing with the detection of B19V using the qPCR methodology in the state of Mato Grosso do Sul, Brazil. The continuous lack of specific studies for B19V in the state of Mato Grosso do Sul may be contributing to the high number of cases that are not laboratory-confirmed (Figure 3). This highlights the need to expand laboratory diagnosis so that B19V can be monitored. However, it is important to point out that the use of a sensitive and specific molecular method is of paramount importance for clarifying the laboratory diagnosis of different viral infections. Thus, our study is the first one showing the circulation of B19V in the state of Mato Grosso do Sul, in the Midwest region of Brazil, using the new protocol developed by F.G. Naveca and his team. Also, our study demonstrates the need and feasibility of implementing B19V detection as a differential diagnosis in patients with acute febrile illness or suspicion of arbovirus infection.

Erythrovirus B19 can occur in individuals of all age groups and from different population groups [26], which is supported by our results (Figure 4A). The B19V infection is usually characterized as acute and self-limited, but clinical manifestations might vary according to the immunological and hematological profile of each person [26]. Some studies describe arthralgia as the most characteristic symptom of B19V infection, especially in adults [27, 28]. Herein, the most common symptoms recorded were, in decreasing order of frequency, fever, myalgia, headache, and, only then, arthralgia (Figure 5). Also, the detection of B19V was significantly associated with the symptoms of “retro-orbital pain”, “leukopenia”, “petechiae” and “malaise” (Figure 4B), thus, diverging from previous studies. However, regardless of the existence of symptoms that might be more associated with B19V infection, there is immense difficulty in distinguishing it from other exanthematous diseases due to the similarity of symptoms. The similarity of the symptoms of erythema infectiosum to other cutaneous exanthematous diseases, together with the wide circulation of arboviruses in Brazil, make the clinical diagnosis a difficult challenge to overcome [6]. In this sense, a study published by [29] showed that about 19% of cases of exanthematous diseases remain without a confirmatory diagnosis; among them, B19V infection is the major cause (33%). Hence, it is essential to carry out a specific laboratory diagnosis for the correct identification of the exanthematous disease agent.

In our study, ten pregnant women tested positive for B19V. Infection by B19V during pregnancy is a serious public health problem, as abortion can occur in the first trimester of pregnancy and in the second trimester, the fetus can develop non-immune hydrops fetalis [30]. The infection can result in vertical transmission to the fetus, causing infection of erythroid precursors and intense hemolysis, leading to severe anemia, fetal hydrops, and death. Rapid correction of anemia by transfusion of packed red blood cells in utero largely prevents fetal death [31]. Unfortunately, it was not possible to track the positive pregnant women to verify the outcome of each case, since they were not diagnosed promptly for adequate follow-up during pregnancy. Women of reproductive age have an annual seroconversion rate of 1.5%, and as in the population group of pregnant women the disease tends to be more severe, so the pregnancy must be monitored medically with the detection of B19V [32].

The identification of B19V “Genotype 1a” (Figure 6) demonstrates the importance of differential laboratory diagnosis using molecular techniques in patients with fever and characteristic symptoms, to assist the health surveillance system through continuous monitoring to detect all circulating genotypes and the spread of the virus in the population. There are three recognized genotypes of B19V (1, 2, and 3), segregated into subtypes 1a, 1b, 2, 3a, and 3b [4, 33–35]. All three genotypes have been reported in both symptomatic and asymptomatic persons, and no association has been established between genotype and clinical manifestations [36–41]. Genotype 1 is the most prevalent in the world. In Brazil, the three genotypes have already been detected, but there is also a predominance of Genotype 1[42, 43]. The first complete genome of B19V genotype 1 a was from a serum sample suspected of dengue infection, from a fatal case of a 12-year-old boy in Rio de Janeiro, Brazil [35]. The viral genome of erythrovirus B19 is highly conserved, with 98-99% similarity between isolates [44, 45], which means that a few sequenced samples are enough to know the circulating genotype.

An important characteristic of B19V infection in Brazil is its cyclic pattern of occurrence every 3 to 5 years, observing years with high infection rates followed by periods with low circulation, as occurred in 1988-1989, 1995-1996, 1999- 2000, 2004-2005, 2009-2010, 2013-2014 [46, 47]. Our results corroborate these previous studies, suggesting that B19V has a cyclical pattern of occurrence in Mato Grosso do Sul (Figure 7). There was a greater number of cases in 2017 and 2018, having decreased in 2019, rising again in 2020, and decreasing again in 2021 and 2022, with a positivity percentage of 20.7%, 13.75%, 8.8 %, 18.3%, 2.3% and 3.4% respectively (Table 1; Figures 3 and 4A). However, further studies are needed over a longer period, since the cycle can last from 4 to 5 years [46]. Additionally, as three years of the study overlapped with the COVID-19 pandemic, this may also have influenced the dynamics of circulation and transmission of B19V.

## CONCLUSION

Timely information represents an essential tool for epidemiological surveillance, as it triggers the “information-decision-action” process, a triad that summarizes the dynamics of its activities, which, as is known, must be initiated from the information of an indication or suspected case of any disease or injury. Although not a new concern, our study demonstrates the importance of the differential diagnosis of B19V in the population, with main attention to pregnant women. As an example, clinical and epidemiological studies carried out in the North region, in 1993, already showed that B19V was becoming a public health problem on the rise at that time [48]. Our study population did not have a laboratory-confirmed diagnosis and, therefore, surveillance did not carry out control actions. This evidences the need for investment in the implementation of methodologies that maintain a high sensitivity of viral detection in areas at risk of emergence of emerging and reemerging pathogens. By identifying the agent that is affecting the population, medical conduct and epidemiological surveillance actions will be better directed. Further, continuous monitoring is needed to detect all known genotypes and the emergence of new genotypes for the control of cases.

## Data Availability

The data that support the findings of this study will be made available in the supplementary of this article.

## Authors’ Contributions

Conceptualization: G.G.C.L, Z.C.F.G, F.G.N, and A.R.M.F; methodology: G.G.C.L, F.G.N, Z.C.F.G, V.A.N, E.F.L, D.M.C.A, and A.R.M.F; investigation: G.G.C.L, Z.C.F.G, V.A.N, D.M.C.A, F.G.N and A.R.M.F; resources: A.R.M.F, Z.C.F.G, C.M.E.C, L.H.F.D and C.C.M.G; data curation: G.G.C.L, V.A.N, E.F.L, F.G.N, D.M.C.A and A.R.M.F; writing – preparation of the original draft: G.G.C.L, A.R.M.F, Z.C.F.G, E.F.L, D.M.C.A, V.A.N, and F.G.N; writing – proofreading and editing: G.G.C.L, Z.C.F.G, A.R.M.F, D.M.C.A, V.A.N, E.F.L, C.M.E.C, L.H.F.D, C.C.M.G and F.G.N; formal analysis: G.G.C.L, Z.C.F.G, A.R.M.F, D.M.C.A, V.A.N and F.G.N; statistical analysis: G.G.C.L and D.M.C.A; supervision: A.R.M.F and Z.C.F.G; financing acquisition: A.R.M.F, Z.C.F.G, C.M.E.C, L.H.F.D and C.C.M.G. All authors read and agreed with the published version of the manuscript.

## Acknowledgments

The authors would like to thank the Central Public Health Laboratory/Mato Grosso do Sul (LACEN) and the Mato Grosso do Sul State Health Secretaria (SES) for their support in carrying out the analyses. The authors also thank FIOCRUZ/MS, FIOCRUZ/AM, FIOCRUZ/RJ, and Evandro Chagas institute for their support and supply of control samples.

## Supplementary Materials

Supplementary 1. Table S1. Primers and probes were used in the study for B19V detection (NAVECA et al., unpublished data).

Supplementary 2. Table S2. Components and volumes used to prepare the reactions for B19V detection (NAVECA et al., unpublished data).

Supplementary 3. Table S3. Primers and probes are used for amplification of the entire genome of B19V.

Supplementary 4. Table S4. Results for 2017 data, with comparison for all 10 orientation models implemented in the R package ‘CircMLE’.

Supplementary 5. Table S5. Results for 2018 data, with comparison for all 10 orientation models implemented in the R package ‘CircMLE’.

Supplementary 6. Table S6. Results for 2019 data, with comparison for all 10 orientation models implemented in the R package ‘CircMLE’.

Supplementary 7. Figure S1. Age group and gender of sampled patients. n, number of patients sampled by age group.

Supplementary 8. Figure S2. The B19V samples that underwent sequencing originate from different municipalities in different geographic areas, demonstrating the wide dissemination of the virus throughout the state.

## Supplementary Material

**Figure S1.**
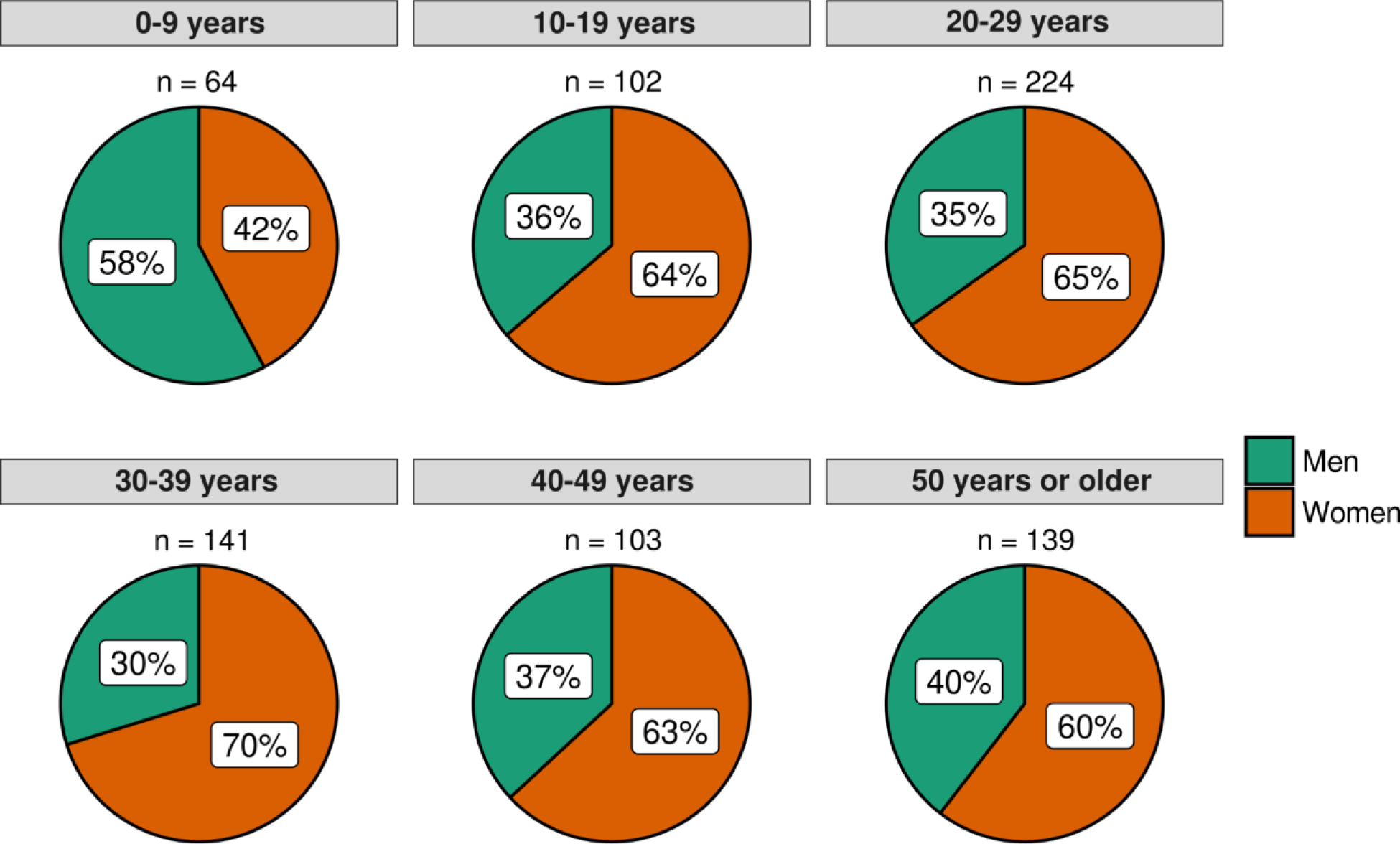
Age group and gender of sampled patients. n, number of patients sampled by age group.

**Figure S2.**
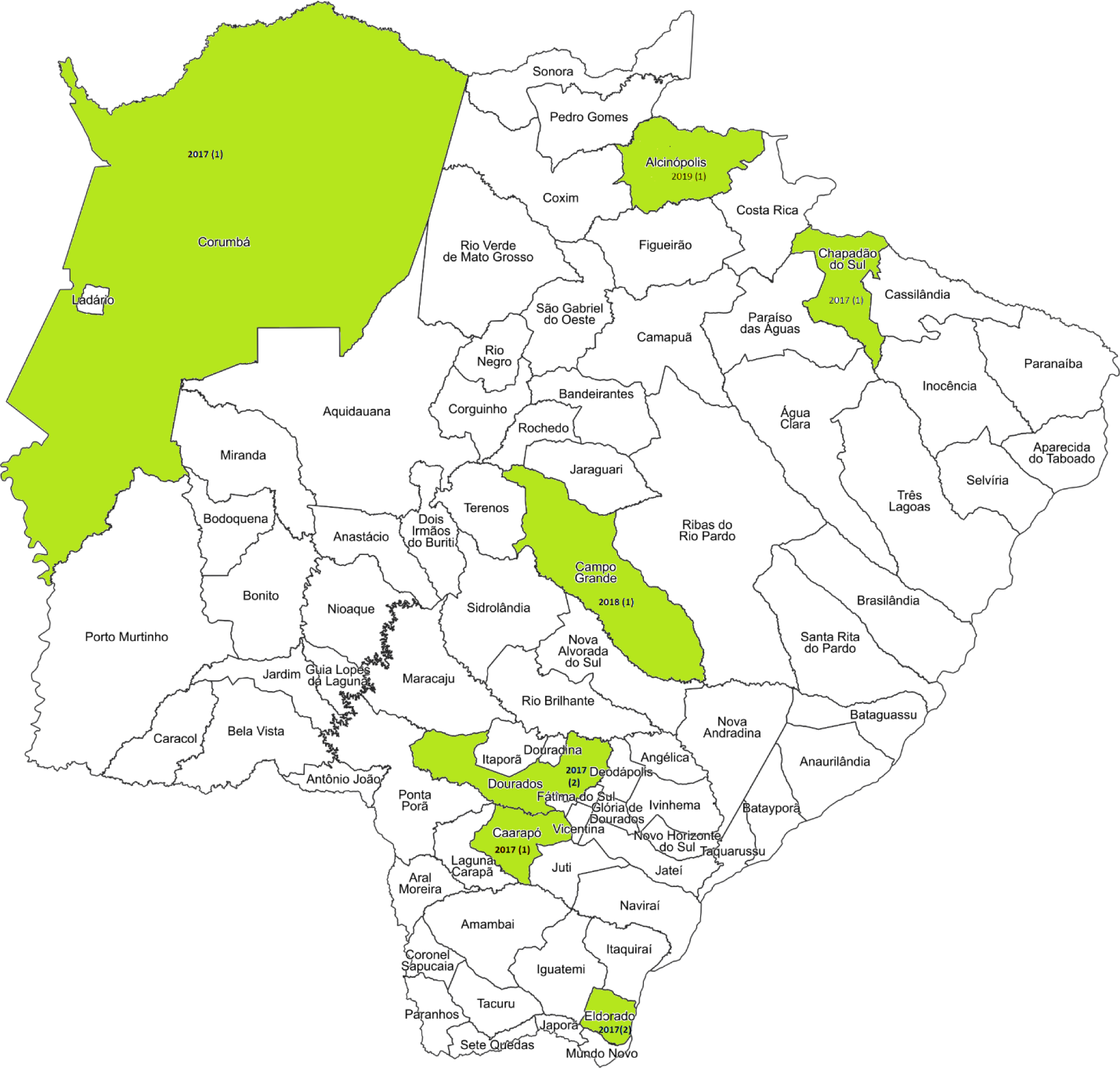
The B19V samples that underwent sequencing originate from different municipalities in different geographic areas, demonstrating the wide dissemination of the virus throughout the state.

**Table S1.**
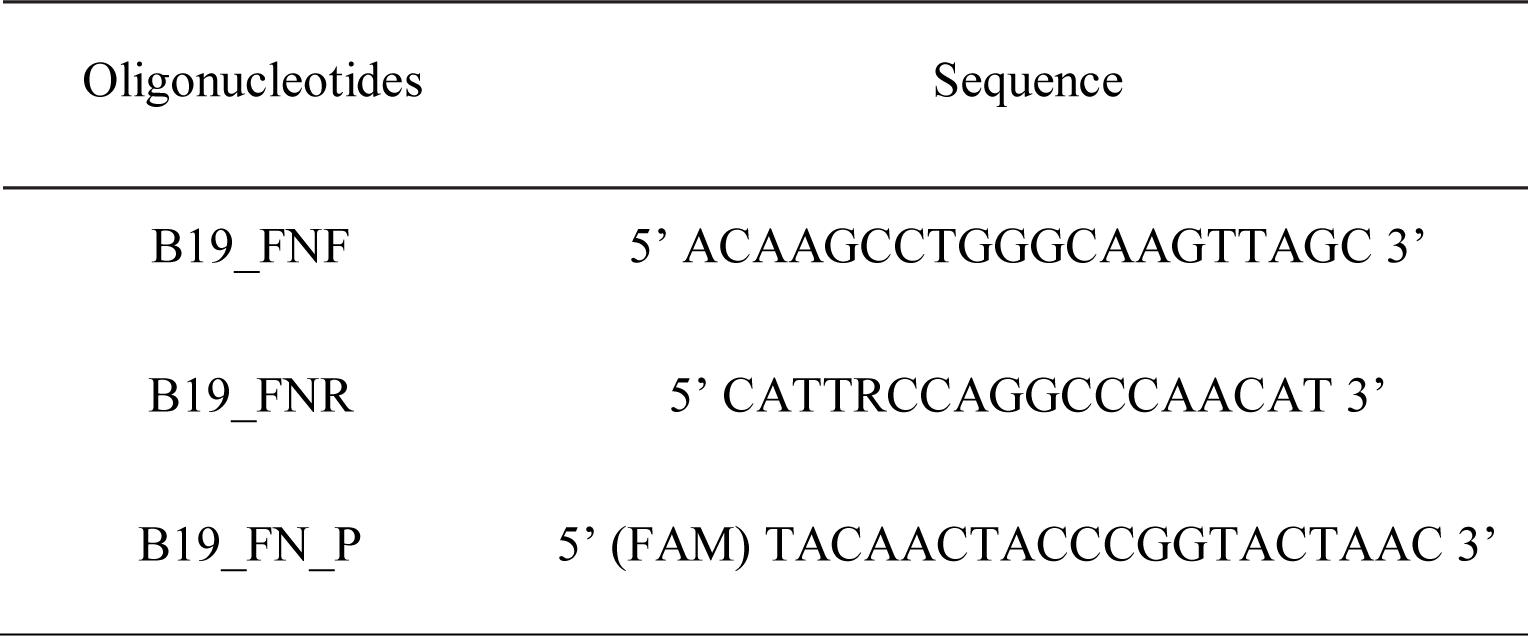
Primers and probes used in the study for B19V detection (NAVECA et al., unpublished data).

**Table S2.** Components and volumes used to prepare the reactions for B19V detection (NAVECA et al., unpublished data).

| Component | 10 µl reaction |
| --- | --- |
| Nuclease-free water | 1.4 µL |
| 2X PCR Master Mix Kapa Probe | 5 µL |
| Primer mix (5 µM) | 0.6 µL |
| Probe (10µM) FAM | 0.1 µL |
| Rox Dye | 0.4 µL |
| Template DNA | 2.5 µL |

**Table S3.**
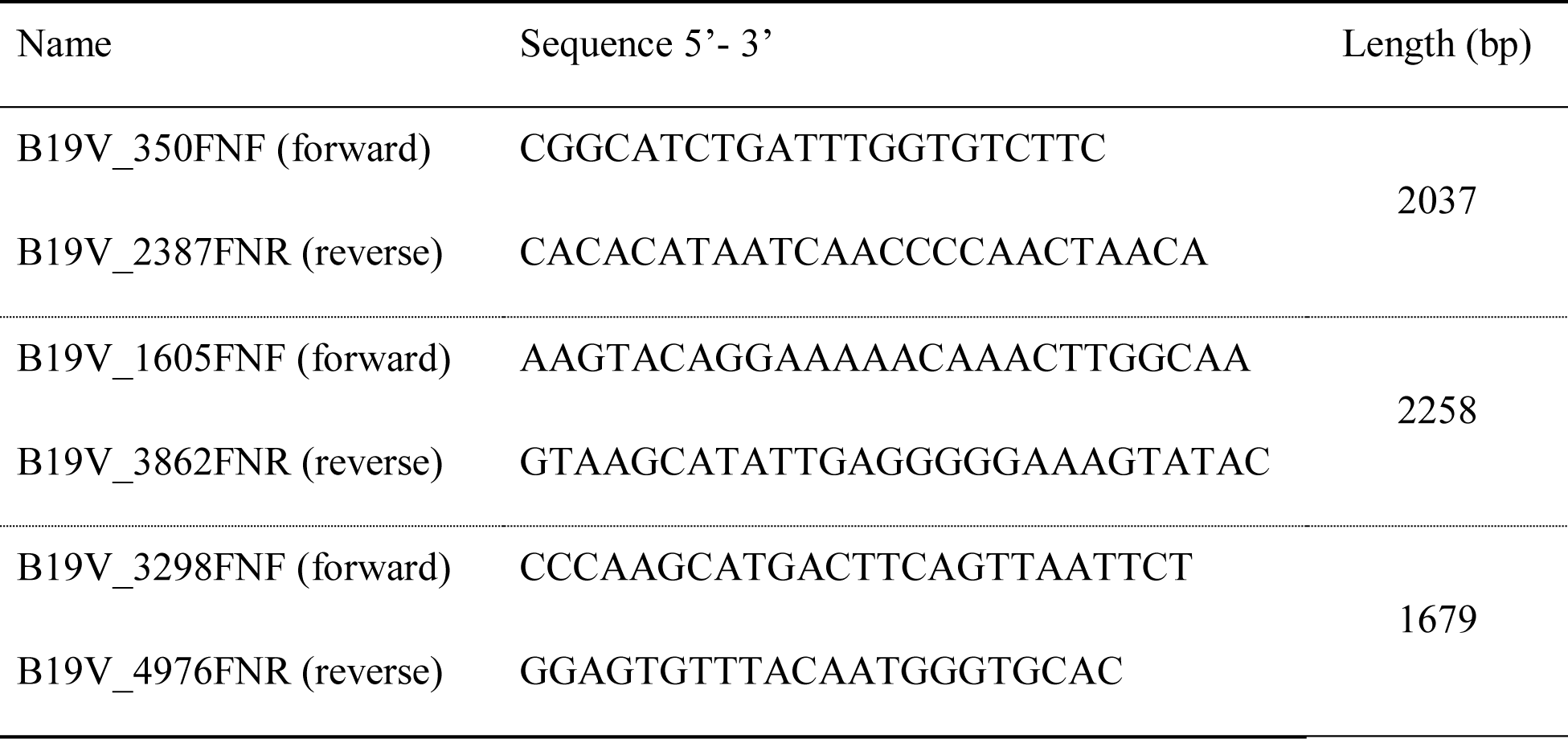
Primers and probes used for amplification of the entire genome of B19V.

**Table S4.** Results for 2017 data, with comparison for all 10 orientation models implemented in the R package ‘CircMLE’. The models are described by five parameters: the mean direction (φ1, in degrees) and concentration parameter (*k*1) for the first mode, the mean direction (φ2, in degrees) and concentration parameter (*k*2) for the second mode, and the proportional size of the first distribution (λ; the second distribution is thus fixed at size 1−λ). They were classified based on Akaike’s Information Criterion with small-sample correction (AICc). ΔAICc, delta AICc; AICc *wi*, model weights; ER, evidence ratio.

| Modelo | $\phi_1$ | $k_1$ | $\lambda$ | $\phi_2$ | $k_2$ | AICc | ΔAICc | AICc $w_i$ | ER |
| --- | --- | --- | --- | --- | --- | --- | --- | --- | --- |
| M3A | 4.40 | 3.18 | 0.50 | 7.54 | 3.18 | 59.11 | 0.00 | 0.38 | - |
| M4A | 4.39 | 3.20 | 0.65 | 7.53 | 3.20 | 60.68 | 1.58 | 0.17 | 2.20 |
| M2C | 4.66 | 49.99 | 0.25 | - | 0.00 | 61.27 | 2.17 | 0.13 | 2.96 |
| M2B | 4.50 | 5.80 | 0.50 | - | 0.00 | 61.60 | 2.50 | 0.11 | 3.49 |
| M3B | 4.45 | 4.05 | 0.50 | 7.59 | 1.64 | 61.81 | 2.70 | 0.10 | 3.86 |
| M1 | - | 0.00 | 1.00 | - | 0.00 | 62.49 | 3.38 | 0.07 | 5.43 |
| M4B | 1.25 | 3.02 | 0.36 | 4.39 | 3.29 | 64.16 | 5.06 | 0.03 | 12.53 |
| M2A | 4.25 | 0.51 | 1.00 | - | 0.00 | 65.23 | 6.13 | 0.02 | 21.40 |
| M5B | 4.36 | 3.31 | 0.64 | 1.31 | 3.02 | 68.20 | 9.10 | 0.00 | 94.38 |
| M5A | 3.45 | 5.00 | 0.41 | 2.66 | 5.00 | 2000000011.33 | 1999999952.23 | 0.00 | - |

**Table S5.** Results for 2018 data, with comparison for all 10 orientation models implemented in the R package ‘CircMLE’. The models are described by five parameters: the mean direction (φ1, in degrees) and concentration parameter (*k*1) for the first mode, the mean direction (φ2, in degrees) and concentration parameter (*k*2) for the second mode, and the proportional size of the first distribution (λ; the second distribution is thus fixed at size 1−λ). They were classified based on Akaike’s Information Criterion with small-sample correction (AICc). ΔAICc, delta AICc; AICc *wi*, model weights; ER, evidence ratio.

| Modelo | $\phi_1$ | $k_1$ | $\lambda$ | $\phi_2$ | $k_2$ | AICc | ΔAICc | AICc $w_i$ | ER |
| --- | --- | --- | --- | --- | --- | --- | --- | --- | --- |
| M5A | 5.08 | 4.62 | 0.47 | 2.49 | 4.62 | 71.17 | 0.00 | 0.38 | - |
| M3A | 5.37 | 3.63 | 0.50 | 8.51 | 3.63 | 71.44 | 0.27 | 0.33 | 1.15 |
| M4A | 2.23 | 3.64 | 0.54 | 5.37 | 3.64 | 74.04 | 2.87 | 0.09 | 4.20 |
| M3B | 2.26 | 4.48 | 0.50 | 5.40 | 2.84 | 74.05 | 2.88 | 0.09 | 4.22 |
| M5B | 2.49 | 4.49 | 0.54 | 5.09 | 4.82 | 74.56 | 3.39 | 0.07 | 5.45 |
| M4B | 2.25 | 4.10 | 0.53 | 5.39 | 3.15 | 77.03 | 5.86 | 0.02 | 18.73 |
| M2C | 2.12 | 30.86 | 0.25 | - | 0.00 | 79.63 | 8.46 | 0.01 | 68.53 |
| M2B | 2.26 | 16.46 | 0.50 | - | 0.00 | 80.36 | 9.19 | 0.00 | 98.94 |
| M1 | - | 0.00 | 1.00 | - | 0.00 | 80.87 | 9.70 | 0.00 | 127.51 |
| M2A | 3.55 | 0.51 | 1.00 | - | 0.00 | 82.80 | 11.63 | 0.00 | 335.68 |

**Table S6.** Results for 2019 data, with comparison for all 10 orientation models implemented in the R package ‘CircMLE’. The models are described by five parameters: the mean direction (φ1, in degrees) and concentration parameter (*k*1) for the first mode, the mean direction (φ2, in degrees) and concentration parameter (*k*2) for the second mode, and the proportional size of the first distribution (λ; the second distribution is thus fixed at size 1−λ). They were classified based on Akaike’s Information Criterion with small-sample correction (AICc). ΔAICc, delta AICc; AICc *wi*, model weights; ER, evidence ratio.

| **Modelo** | **$\phi_1$** | **$k_1$** | **$\lambda$** | **$\phi_2$** | **$k_2$** | **AICc** | **ΔAICc** | **AICc $w_i$** | **ER** |
| --- | --- | --- | --- | --- | --- | --- | --- | --- | --- |
| M2A | 1.31 | 1.46 | 1.00 | - | 0.00 | 77.34 | 0.00 | 0.72 | - |
| M5A | 0.33 | 2.58 | 0.37 | 1.82 | 2.58 | 81.43 | 4.09 | 0.09 | 7.73 |
| M2C | 1.33 | 2.15 | 0.75 | - | 0.00 | 81.65 | 4.32 | 0.08 | 8.66 |
| M2B | 1.39 | 2.36 | 0.50 | - | 0.00 | 82.11 | 4.78 | 0.07 | 10.89 |
| M3B | 4.62 | 0.00 | 0.50 | 7.76 | 2.24 | 84.84 | 7.51 | 0.02 | 42.64 |
| M4B | 4.40 | 0.00 | 0.27 | 7.54 | 2.30 | 85.00 | 7.67 | 0.02 | 46.20 |
| M5B | 6.28 | 2.53 | 0.49 | 1.71 | 3.82 | 88.08 | 10.74 | 0.00 | 215.33 |
| M1 | - | 0.00 | 1.00 | - | 0.00 | 91.89 | 14.56 | 0.00 | 1450.15 |
| M3A | 1.52 | 0.98 | 0.50 | 4.66 | 0.98 | 95.82 | 18.48 | 0.00 | 10315.06 |
| M4A | 5.56 | 0.81 | 0.25 | 8.70 | 0.81 | 96.28 | 18.95 | 0.00 | 13008.91 |

## References

1. YE, C., et al., *Parvovirus-like particles in human sera.* Lancet (London, England), 1975. 1(7898).

2. Moore, T.L.M., Parvovirus-associated arthritis: Current Opinion in Rheumatology. 2023.

3. Cotmore, S.F., et al., The family Parvoviridae. Archives of Virology, 2013. 159(5): p. 1239–1247.

4. A, S., et al., Genetic diversity within human erythroviruses: identification of three genotypes. Journal of Virology, 2002. 76(18).

5. A, d.S.C., et al., Detection of the human parvovirus B19 in a blood donor plasma in Rio de Janeiro. Memorias do Instituto Oswaldo Cruz, 2023. 84(2): p. 279–280.

6. Cnc Garcia, R. and L.A. Leon, Human parvovirus B19: a review of clinical and epidemiological aspects in Brazil. Future Microbiol, 2021. 16(1): p. 37–50.

7. Alves, A. Beyond arboviruses: A multicenter study to evaluate the differential diagnosis of rash diseases and acute febrile illness cases in Rio de Janeiro, Brazil | PLOS ONE. 2023; Available from: https://journals.plos.org/plosone/article?id=10.1371/journalpone.0271758.

8. RC, C., et al., Human parvovirus B19 infections among exanthematic diseases notified as measles. Revista da Sociedade Brasileira de Medicina Tropical, 1997. 30(1).

9. Anisimova, M., et al., Approximate Likelihood-Ratio Test for Branches: A Fast, Accurate, and Powerful Alternative. Systematic Biology, 2006. 55(4): p. 539–552.

10. Makowski, CRAN - Package jtools. 2023.

11. Lüdecke, D., et al., *performance: An R Package for Assessment*, Comparison and Testing of Statistical Models. Journal of Open Source Software, 2021. 6(60): p. 3139.

12. Bates, D., et al., Fitting Linear Mixed-Effects Models using lme4. 2014.

13. Team, R.C. R: The R Project for Statistical Computing. 2022; Available from: https://www.r-project.org/.

14. Long, CRAN - Package jtools. 2023.

15. Harting, CRAN - Package DHARMa. 2023.

16. Almeida, A., C. Codeço, and P.M. Luz, Seasonal dynamics of influenza in Brazil: the latitude effect. BMC Infectious Diseases, 2018. 18(1): p. 1–9.

17. L, L., R. GD, and M. EP, The Hermans-Rasson test as a powerful alternative to the Rayleigh test for circular statistics in biology. BMC Ecology, 2019. 19(1).

18. Landler, L., et al., Model selection versus traditional hypothesis testing in circular statistics: a simulation study. Biology Open, 2020. 9(6).

19. RR, F. and J. S, Bringing the analysis of animal orientation data full circle: model-based approaches with maximum likelihood. The Journal of Experimental Biology, 2017. 220(Pt 21).

20. Schnute, J., Statistical analysis of animal orientation data. 1992.

21. Ojeda, V., et al., Latitude does not influence cavity entrance orientation of South American avian excavators. Ornithology, 2022. 138(1).

22. Burnham, K.P., D.R. Anderson, and K.P. Huyvaert, AIC model selection and multimodel inference in behavioral ecology: some background, observations, and comparisons. Behavioral Ecology and Sociobiology, 2010. 65(1): p. 23–35.

23. S, P., *A primer on model selection using the Akaike Information Criterion*. Infectious Disease Modelling, 2020. 5.

24. Agostineli, C. *R-Forge: Circular Statistics: Project Home*. 2023; Available from: https://r-forge.r-project.org/projects/circular/.

25. E, M., et al., *Laboratory assessment and diagnosis of congenital viral infections: Rubella, cytomegalovirus (CMV), varicella-zoster virus (VZV), herpes simplex virus (HSV), parvovirus B19 and human immunodeficiency virus (HIV).* Reproductive toxicology (Elmsford, N.Y.), 2006. 21(4).

26. Qiu, J., M. Söderlund-Venermo, and N.S. Young, Human Parvoviruses. 2016.

27. Suzuki, M., et al., Parvovirus B19 Infection: A Vasculitis Masquerade in an Elderly Patient. American Journal of Case Reports, 2023. 24.

28. Young, N.S. and K.E. Brown, Parvovirus B19. 10.1056/NEJMra030840, 2004.

29. T, V.A., et al., [Incidence and clinical characteristics of maculopapular exanthemas of viral etiology]. Atencion primaria, 2003. 32(9).

30. Bouraddane, M., K. Warda, and S. Zouhair, Parvovirus B19, and Pregnant Women: A Bibliographic Review. Open Journal of Obstetrics and Gynecology, 2021. 11(11): p. 1543.

31. SN, S., et al., *Molecular and phylogenetic analyses of human Parvovirus B19 isolated from Brazilian patients with sickle cell disease and* β*-thalassemia major and healthy blood donors*. Journal of Medical Virology, 2012. 84(10).

32. WC, K. and A. SP, Human parvovirus B19 infections in women of childbearing age and within families. The Pediatric Infectious Disease Journal, 2023. 8(2): p. 83–87.

33. Toan, N.L., et al., Phylogenetic analysis of human parvovirus B19, indicating two subgroups of genotype 1 in Vietnamese patients. 2006.

34. Parsyan, A., et al., Identification and genetic diversity of two human parvovirus B19 genotype 3 subtypes. 2007.

35. Conteville, L.C., et al., Parvovirus B19 1A complete genome from a fatal case in Brazil. Memórias do Instituto Oswaldo Cruz, 2023. 110: p. 820–821.

36. Sanabani, S., et al., Sequence Variability of Human Erythroviruses Present in Bone Marrow of Brazilian Patients with Various Parvovirus B19-Related Hematological Symptoms. 2006.

37. MW, M.-d.B., et al., Global co-existence of two evolutionary lineages of parvovirus B19 1a, different in genome-wide synonymous positions. PloS One, 2012. 7(8).

38. Rahiala, J., et al., Human parvoviruses B19, PARV4 and bocavirus in pediatric patients with allogeneic hematopoietic SCT. Bone Marrow Transplantation, 2013. 48(10): p. 1308-1312.

39. Oliveira, M.I.d., et al., Genotype 1 of human parvovirus B19 in clinical cases. Revista da Associação Médica Brasileira, 2017. 63: p. 224–228.

40. Seetha, D., et al., Molecular-genetic characterization of human parvovirus B19 prevalent in Kerala State, India. Virology Journal, 2021. 18(1): p. 1–8.

41. M, H., et al., Parvovirus B19 infection in pediatric allogeneic hematopoietic cell transplantation - Single-center experience and review. Transplant infectious disease: an official journal of the Transplantation Society, 2023. 25(2).

42. RB, F., et al., Molecular characterization of human erythrovirus B19 strains obtained from patients with several clinical presentations in the Amazon region of Brazil. Journal of Clinical Virology: the official publication of the Pan American Society for Clinical Virology, 2008. 43(1).

43. RC, C.G., et al., Molecular diversity of human parvovirus B19 during two outbreaks of erythema infectiosum in Brazil. The Brazilian journal of infectious diseases: an official publication of the Brazilian Society of Infectious Diseases, 2017. 21(1).

44. Hicks, K.E., et al., Sequence analysis of a parvovirus B19 isolate and baculovirus expression of the non-structural protein. Archives of Virology, 2023. 141(7): p. 1319–1327.

45. Cubel, R.C.N., et al., Human parvovirus B19 infections among exanthematic diseases notified as measles. Revista da Sociedade Brasileira de Medicina Tropical, 1997. 30:p. 15–20.

46. Oliveira, M.I.d., et al., Genotype 1 of human parvovirus B19 in clinical cases. Revista da Associação Médica Brasileira, 2023. 63: p. 224–228.

47. Garcia, R.d.C. and L.A. Leon, Human parvovirus B19: a review of clinical and epidemiological aspects in Brazil. 10.2217/fmb-2020-0123, 2021.

48. Freitas, R.B.d., et al., Parvovirus B19 antibodies in sera of patients with unexplained exanthemata from Belém, Pará, Brazil. 1993.

